# Multi-ancestry genome-wide association study of neutrophil-lymphocyte ratio and polygenic risk score development to explore causal association with diabetic retinopathy

**DOI:** 10.1101/2024.06.19.24309194

**Authors:** Aravind Lathika Rajendrakumar, Anand Thakarakkattil Narayanan Nair, Mehul Kumar Chourasia, Charvi Nangia, Sundararajan Srinivasan, Venkateshan Radha, Ranjit Mohan Anjana, Moneeza K Siddiqui, Weihua Meng, Viswanathan Mohan, Colin N A Palmer

## Abstract

**Background:** Neutrophil–lymphocyte Ratio (NLR) is a biomarker of inflammation and was associated with diabetic retinopathy (DR) in earlier studies.

**Objective:** To investigate the genetic loci influencing NLR and to estimate the heritability and causality of DR with the NLR polygenic risk score (PRS).

**Design:** Genome-wide association study, conditional analysis, Fine and Gray model (FGR), Mendelian Randomization (MR)

**Setting:** Scottish and South Indian populations drawn from population cohorts and electronic medical records.

**Participants:** 29,317 individuals, with a considerable proportion diagnosed with diabetes.

**Measurements:** Effect estimates from GWAS to compute PRS and causal association with DR.

**Results:** Heritability estimates for the Scottish and Indian cohorts were 35.3% and 8.7% respectively. The top Single Nucleotide Polymorphisms (SNPs) in the multi-ancestry analysis (n=29,317) were intergenic: rs1825819 (Chr4:T/C) (Beta=-0.05, p=2.00×10^-9^), rs2980871 (Chr8:A/G) (Beta=0.04, p=4.64×10^-8^), rs2227322 (Chr17:C/G) (Beta=0.07, p=4.12×10^-20^) and rs4808047 (Chr19:T/C) (Beta= - 0.07, p=5.93×10^-12^). For the construction of best-fit PRS, we used 74,377 of 55,333,12 variants. There was a dose-response relationship between the PRS and NLR. The subhazard ratio (sHR) for NLR PRS association with DR was not statistically significant sHR=1.01 (95% CI: 0.97, 1.06, p=0.48). Null associations were observed in both cross-sectional and time-based MR analyses for PRS with DR.

**Limitations:** A substantial proportion of the dataset was used for training the PRS algorithm. Due to trans-ancestry differences, PRS and subsequent analysis were conducted only in the Scottish cohorts.

**Conclusions:** Multiple novel intergenic SNP associations were discovered, complementing those previously identified. Of these, some SNPs were also associated with genes known to regulate white blood cells, but not specifically NLR. More studies are required to confirm the causality between systemic inflammation and DR.

**Primary Funding Source:** National Institute for Health Research, Pioneer and Leading Goose R&D Program of Zhejiang 2023, and the Ningbo International Collaboration Program 2023.

## Introduction

Microvascular abnormalities are not observed in certain individuals despite having long-term diabetes but the exact reasons for this are ill understood.^1–4^ In complex diseases, the genetic liability of an individual, can be summarised into an informative risk score that can be used in disease prediction models.^5,6^ Diabetes and inflammation share overlapping biological mechanisms.^7^ The global burden of diabetic retinopathy (DR) was close to 23% and more than 6% experience vision problems.^8^ Increased concentrations of several inflammatory molecules were noted in tissues and vitreous fluids of individuals with DR.^9^ Genetic modification of specific inflammatory pathways was shown to arrest the progress of DR.^10^ Leukocyte recruitment in DR was found to prolong the inflammatory activity.^11^ Neutrophil–Lymphocyte Ratio (NLR) was shown to be more clinically informative for several clinical conditions, than a more established marker of inflammation such as C-reactive protein (CRP) in interferonopathies.^12^ Hence, modeling the joint variations in neutrophils and lymphocytes could be more informative than using them individually as markers of inflammation.^13–17^ Indeed, the NLR has been shown to be associated with diabetes and its complications.^18–21^ However, no studies have directly investigated the genetic causality of the observed associations. We aimed to identify new genetic loci for NLR with a strict phenotype definition. An allele score for NLR could be more powerful as it entails comprehensive information from multiple genetic loci. We hypothesized that the weighted NLR allele score could be used as an ideal instrumental variable to represent the genetic propensity for a heightened inflammatory response. We therefore performed a genome-wide association study (GWAS) for NLR and used the resulting PRS, both as an exposure and instrument variable to predict and find causal association with DR.

## Methods

### Cohort description and sample selection

The India-Scotland Partnership for Precision Medicine in Diabetes (INSPIRED) is a collaboration between Dr. Mohan’s Diabetes Specialties Centre (DMDSC), and the associated Madras Diabetes Research Foundation (MDRF) at Chennai in South India and the University of Dundee, Scotland.In the discovery phase, we meta-analyzed 3 Scottish cohorts: Genetics of Diabetes Audit and Research in Tayside Scotland (GoDARTS-T2D), Scottish Health Research Register (SHARE), and GoDARTS non-type 2 diabetes controls (GoDARTS controls. However, a small proportion of GoDARTS controls developed diabetes after enrollment. These Scottish cohorts consist of individuals of predominantly white European ancestry. These cohorts and the data linkage strategies used for research have been described previously.^22,23^ The findings were replicated in the DMDSC cohort 1 and cohort 2. The main difference between the DMDSC cohorts was the sampling strategy employed for genotyping. The participants were selected retrospectively in DMDSC cohort 1. NLR PRS was computed in SHARE using GWAS estimates from the GoDARTS T2D and controls.

### Genotyping

Genotyping methods for the Scottish and Indian cohorts have been described previously.^24^ The DNA was extracted from the stored blood plasma. A trained bioinformatician performed the quality control using standard protocols. For all the cohorts, Haplotype Reference Consortium (HRC) was used to infer the missing genotypes. We utilized the GRCh37 assembly (Genome Research Consortium human build 37) to locate SNPs. Affymetrix 6.0, Illumina Human Omni Express, and Broad Institute platforms were used for genotyping GoDARTS. Both, DMDSC and SHARE participants were assayed using the Illumina GSA platform. Alleles with a minimum allele frequency (MAF) greater than 0.05 were considered for GWAS. Additionally, we excluded single nucleotide polymorphism (SNPs) with an info score >0.70 and Hardy-Weinberg equilibrium (HWE) <1×10^-10^. Principal components were computed using Plink Version 1.9.^25,26^ We have used STREGA guidelines to report our findings.^27^

### Definition of NLR and DR phenotypes

The Scottish diabetic retinopathy grading scheme was used to ascertain DR status (yes/no) in the Scottish cohort.^28^ Details of retinal screening in Tayside were described previously.^29^ For the time-based Mendelian Randomization (MR) analysis, the first reported DR (any grade from R1 to R4) was considered. For non-DR individuals, the maximum follow-up date was considered as the censoring date. NLR was calculated as the absolute count of neutrophils divided by the absolute count of lymphocytes from the hematology dataset. All values of neutrophils and lymphocytes above and below 5 standard deviations (SD) were removed. The change in NLR above 2.5 from the previous reading was considered and hence excluded. For, individuals having a single NLR reading, values above 5 were excluded. NLR values after diagnosis or treatment for malignancies or within a window period of 31 days after the diagnosis of infectious diseases were discarded. We also did not retain an NLR reading within 28 days after the first reading, assuming that as a case of possible visit for a disease condition.

### Ethics Approval

#### Tayside, Scotland UK

Ethical approval for the study was provided by the Tayside Medical Ethics Committee (REF:053/04) and the study has been carried out in accordance with the Declaration of Helsinki.

#### SHARE, Scotland UK

Ethical approval of the study was provided by the ethics committee SHARE East of Scotland (REF: NHS REC 13/ES/0020).

#### DMDSC

NIHR Global Health Research Unit on Global Diabetes Outcomes Research, Institutional Ethics Committee of Madras Diabetes Research Foundation, Chennai, India. IRB number IRB00002640. Granted 24^th^ August 2017.

## Statistical Analysis

### GWAS analysis and ascertainment of independent loci

The analytical approaches are shown in Supplementary Figure 1. Continuous variables were summarized as mean/median and categorical variables as frequencies and percentages. GWAS was performed in SNPtest version 2.5.2 with 10 principal components, age, sex, and diabetes status as covariates. Genome-wide Complex Trait Analysis (GCTA) software was used to identify independent genomic loci and genomic inflation was controlled during the computation process.^30^ We used the cojo-joint function in the GCTA suite which uses all the SNPs to calculate joint effect size without eliminating less important SNPs from the model. GWAS summary estimates were meta-analyzed with GWAMA (Genome-Wide Association Meta-Analysis) by applying the the genomic control option (-gco) in GWAMA to address inflation and assuming an inverse variance weighted model with a p value significance cut off of 5×10^-8.31^

### Gene enrichment, functional analysis, heritability estimation, and gene network analysis

Functional Mapping and Annotation of Genome-wide Association Studies (FUMA) was used to create the regional plot, annotation, enrichment statistics, and gene-based tests. We also took annotations from gnomAD (https://gnomad.broadinstitute.org/), UCSC browsers (https://genome.ucsc.edu/), and genecards (https://www.genecards.org/) if the annotations were not available in the FUMA. GeneMANIA was used to understand the co-expression and pathway analysis (https://genemania.org). GeneMANIA predicts weighted gene networks based on well-known genetic databases to visualize gene interactions.^32^ Narrow-sense heritability (h^2^ was estimated using LDAK software.^33^

### PRS estimation

PRSice-2 and packages in R version 3.6.1 (The R Foundation for Statistical Computing, Vienna, Austria) were used to generate PRS and perform competing risks analyses, respectively.^34–36^ The Clumping and Thresholding method (C+T) in PRSice-2 was utilized for SNP thinning. GoDARTS T2D and GoDARTS controls were used as the base data and SHARE as the target data.

### Competing risks regression and MR analysis

MR and competing risk analyses were performed using allele score as an instrument in the SHARE cohort. Fine and Gray analysis using the riskRegression package with deaths as a competing event adjusting for age, sex, HbA1c, BMI, and creatinine.^37,38^ Subsequently, The PRS from the SHARE cohort was used to test the causal relationship between NLR and DR using the one-sample Mendelian randomization (MR) method. For the Mendelian randomization analysis, we experimented with both cross-sectional and time-based models for the same DR phenotypes using R codes, and the *ivtools* package was used for survival MR.^39^

## Results

### Baseline characteristics of the participants

The characteristics of the Scottish discovery cohort (white Europeans, n= 21,153) are shown in Supplementary Table 1. Among these, the GoDARTS had the highest sample size. Over 96% of individuals had diabetes in the Affymetrix and Illumina sub-cohorts. The median NLR for all the Scottish cohorts was above 2. GoDARTS controls had the lowest median NLR (2.04), and only 7.7% had diabetes. Participants in the GoDARTS were older in comparison to the SHARE cohort. Except for GoDARTS controls, the proportion of males in all other cohorts was above 50%. In the DMDSC (Asian Indians = 8,164), the median NLR (1.93) and the median age was lower (less than 60 years) than the Scottish cohort (Supplementary Table 2).

### Discovery Phase in white European cohorts

Meta-analysis of the 3 Scottish cohorts revealed 150 significant SNPs, most of which were in Chromosome 4 and Chromosome 17 (Supplementary Figures 2-3). Out of these, 128 were from Chromosome 17 and were in high linkage disequilibrium (LD). Only 3 SNPs were independently associated with the NLR trait, of which only one, rs3826331 in chromosome 17, and the rest, rs6841652 and rs16850400 in chromosome 4 (Table 1). Among these, rs3826331 is an intron near the neutrophil gene *PSMD3*, rs6841652 can be mapped either to the *LINCO2513* or *TBC1D1* genetic regions, and the gene corresponding to rs16850400 is yet to be determined. Of the 3 SNPs, the association signal for rs16850400 was suggestive (p=2.39 x 10^-6^). The estimated SNP heritability was 35.3% (SD: 0.03). The genomic inflation factor (λ) was within reasonable limits for individual λ GWAS but was slightly on the higher side for the fixed effects meta-analysis (λ =1.09).

**Table 1.**
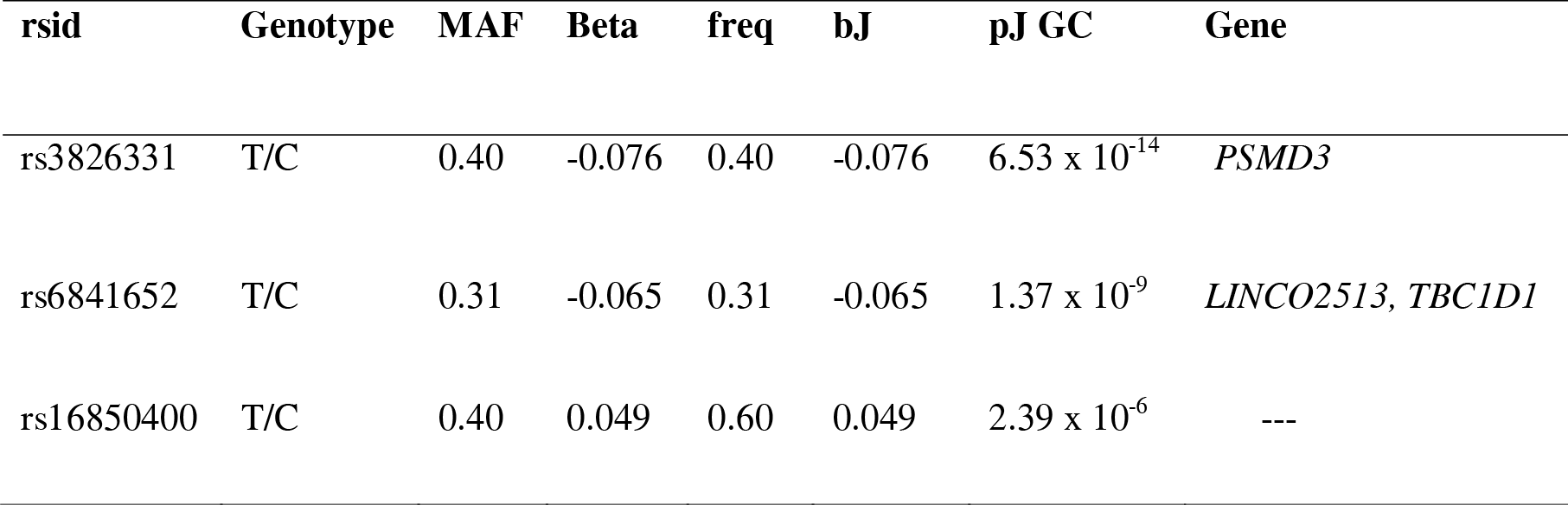
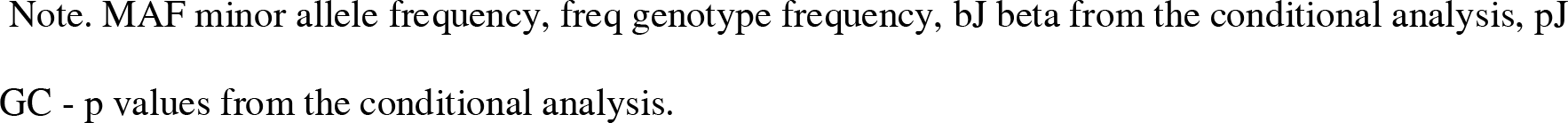
SNP estimates were identified as conditionally independent using GCTA in the Scottish GWAS meta-analysis.

| rsid | Genotype | MAF | Beta | freq | bJ | pJ GC | Gene |
| --- | --- | --- | --- | --- | --- | --- | --- |
| rs3826331 | T/C | 0.40 | -0.076 | 0.40 | -0.076 | $6.53 \times 10^{-14}$ | *PSMD3* |
| rs6841652 | T/C | 0.31 | -0.065 | 0.31 | -0.065 | $1.37 \times 10^{-9}$ | *LINCO2513, TBC1D1* |
| rs16850400 | T/C | 0.40 | 0.049 | 0.60 | 0.049 | $2.39 \times 10^{-6}$ | --- |

Note. MAF minor allele frequency, freq genotype frequency, bJ beta from the conditional analysis, pJ GC - p values from the conditional analysis.

### Replication phase in Asian Indian cohort

The DMSDC GWAS plots are shown in Supplementary Figures 5 and 6. The genome-wide search for SNPs in the DMDSC cohorts found two highly correlated SNPs in Chromosome 6. Table 2 shows the details of these SNPs: rs17330192 (p=1.8 x 10^-8^), and rs139801819 (p=2.4 x10^-8^) were introns found near the non-coding region of *LOC102724591* (the RNA gene). Heritability estimated in the Indian population was 8.7% (SD: 0.07). During the replication effort, a reversal of SNP effects was noted (Table 3), and rs3826331 (T/C) remained statistically significant but with a reversed effect as a trait-increasing allele (Beta =0.06, p=1.5 x 10^-6^). We were unable to replicate rs6841652 (p=0.06) and rs16850400 (p=0.08) in the DMDSC cohort. Arguably, the MAF was relatively common in both ancestries and therefore would not have been the reason for non-replication.

**Table 2.**
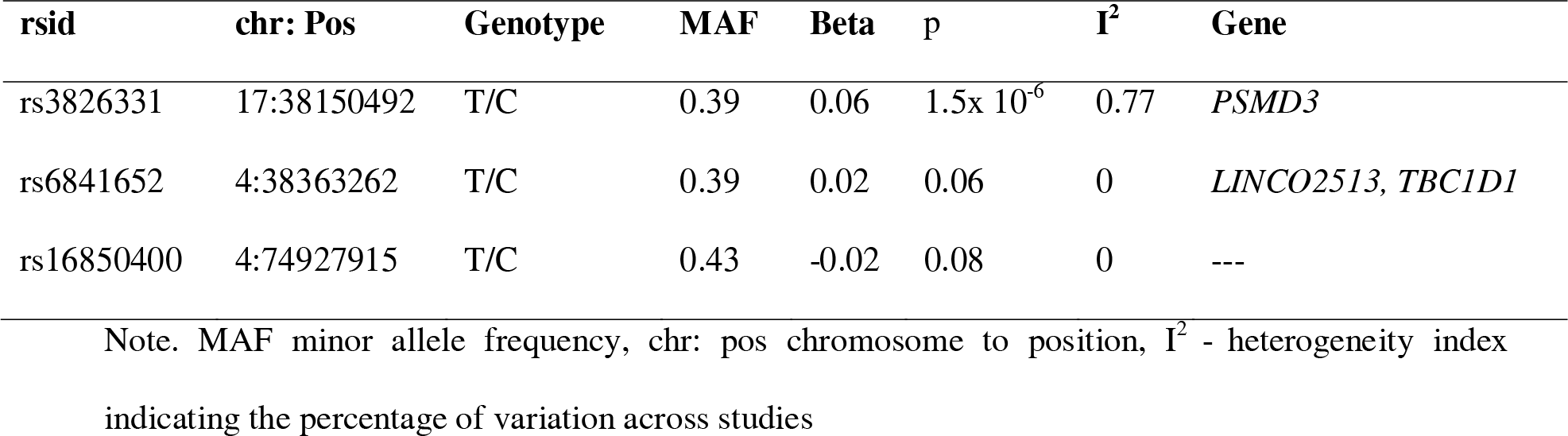
Estimates of SNP associations with NLR from the replication analysis in the DMDSC (n=8,164)

| rsid | chr: Pos | Genotype | MAF | Beta | p | I<sup>2</sup> | Gene |
| --- | --- | --- | --- | --- | --- | --- | --- |
| rs3826331 | 17:38150492 | T/C | 0.39 | 0.06 | $1.5 \times 10^{-6}$ | 0.77 | *PSMD3* |
| rs6841652 | 4:38363262 | T/C | 0.39 | 0.02 | 0.06 | 0 | *LINCO2513, TBC1D1* |
| rs16850400 | 4:74927915 | T/C | 0.43 | -0.02 | 0.08 | 0 | --- |

Note. MAF minor allele frequency, chr: pos chromosome to position, I<sup>2</sup> - heterogeneity index indicating the percentage of variation across studies

**Table 3.**
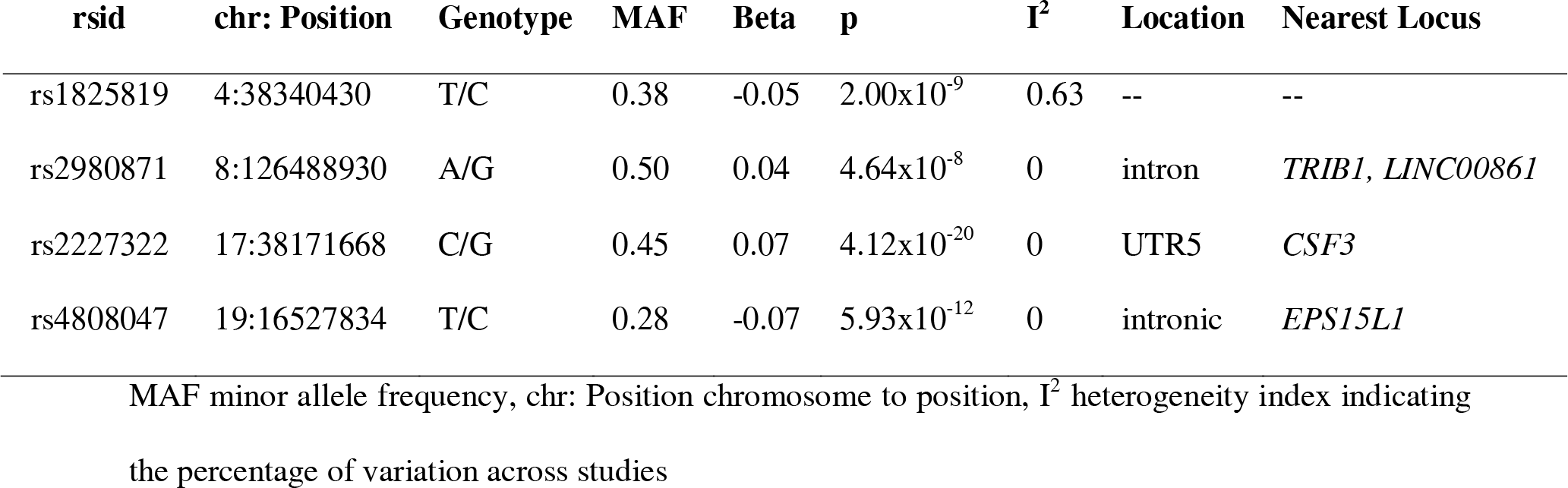
Lead SNPs identified from trans-ancestry inverse variance meta-analysis of Scottish and Indian Cohorts (n=29,317)

### Fixed effects trans-ancestry meta-analysis phase

The final sample size for the FE trans-ancestry meta-analysis was 29,317, and the SNP associations are shown in Supplementary Figures 6-7. We identified two novel SNPs in Chromosome 4 and 8: rs1825819 (p=2.0×10^-9^) and rs2980871 (p=4.6×10^-8^) respectively (Table 3). The genomic inflation factor post-correction was 1.07. Regional plots show that both chromosome 17 and chromosome 19 had many SNPs in high linkage disequilibrium (Supplementary Figures 8-12). Conversely, chromosome 8 and chromosome 4 had very few highly correlated SNPs.

In the FE trans-ancestry analysis, all the lead SNPs were found to be in the non-coding regions. Four lead SNPs were found statistically to be significant, of which rs2227322 had the highest trait-increasing effect size (0.07, p=4.12×10^-20^). This UTR5 variant was located on chromosome 17 in the *CSF3* gene. This gene was responsible for granulocyte synthesis and also cytokine-related inflammatory responses (https://www.ncbi.nlm.nih.gov/gene/1440). The second notable SNP was rs4808047 on Chromosome 19 (-0.07, p=5.93×10^-12^) in the *EPS15L1* gene and had the lowest MAF (0.28) among the lead SNPs. SNP rs2980871 in Tribbles homolog 1 (*TRIB1*) was another interesting finding. In mouse models, *TRIB1* expression affects hepatic lipid synthesis and glycogenesis.^40^ On chromosome 4, rs1825819 with a MAF of 0.38 was found to be associated with NLR (-0.05, 2.00×10^-9^). However, there were no known previous phenotype associations or functions identified for this specific SNP, making it a novel locus for NLR. The sentinel SNPs identified in the trans-ancestry meta-analysis generally showed a consistent direction in terms of their effects, except for the SNP rs1825819. The direction of this SNP was trait-increasing in the Broad array and reversed for other cohorts.

Figure 1 shows the significant genes in the trans-ancestry meta-analysis, in the gene-based test, *CLECL1* (chromosome 12), *GSDMA* (chromosome 17)*, CSF3* (chromosome 17)*, PSMD3* (chromosome 17)*, MED24* (chromosome 17)*, THRA* (chromosome 17)*, CTD-3222D19.2* (chromosome 19)*, EPS15L1* (chromosome 19)*, CALR3* (chromosome 19)*, CHERP* (chromosome 19)*, CTC429P9.4* (chromosome 19)*, MED26* (chromosome 19), and *SLC35E1* (chromosome 19) were identified. *CLECL1* on chromosome 12 was another important gene that was identified. Among the 13 genes reported in the gene-based test, *GSDMA, CLECL1, CALR3,* and *SLC35E1* did not have any functional relationships with other genes in the network analysis. No gene interaction or pathway information was available for the genes *CTD-3222D19.2* and *CTC429P9*.

**Figure 1.**
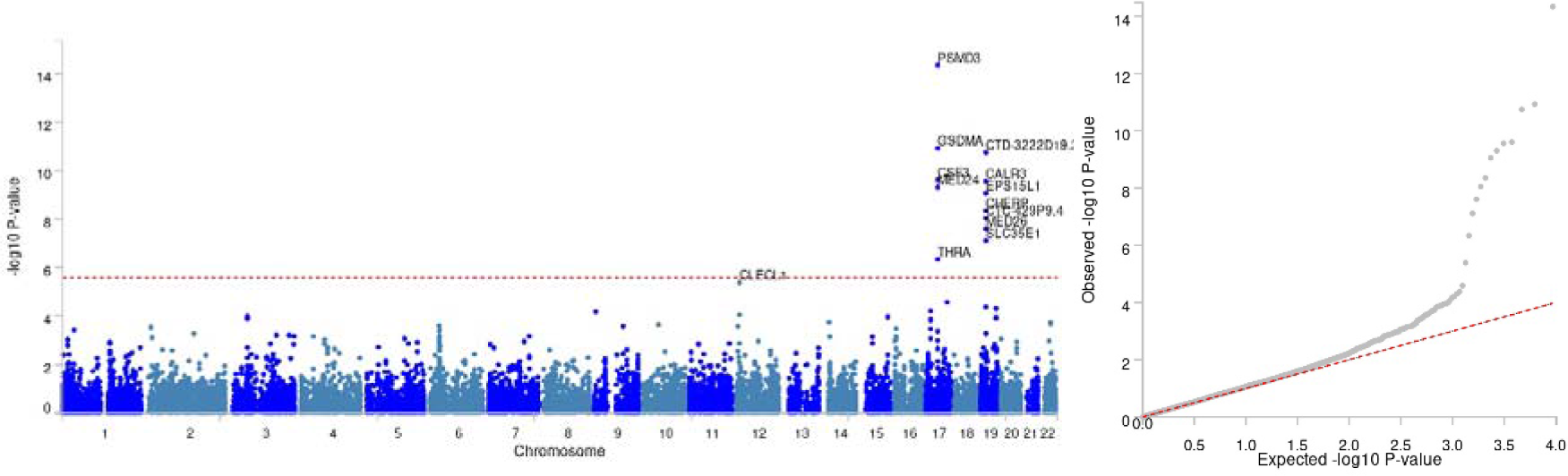
Manhattan plot and (left) Q-Q plot (right) corresponding to the gene set analysis from the trans-ancestry GWAS summary (n= 29,317) Note: The SNPs corresponded to 9528 protein coding genes, the genome-wide significance threshold (red dashed line in the plot) was established at P = 0.05/9528 = 5.248e-6.

The independent SNPs identified in the discovery set also remained statistically significant in the FE tran-ancestry analysis. Supplementary Figure 13 illustrates the details of the number of SNPs and genes in the important genomic loci in the meta-analysis. Tissue-wise differential gene expression analysis showed that the genes were generally upregulated in the adrenal glands, whole blood, and multiple regions of the brain (Supplementary Figure 14). However, these did not surpass the statistical significance threshold. Gene expression for nine selected significant genes from the gene-based analysis is plotted in Supplementary Figure 15. *PSMD3*, the major gene in our analysis, along with *MED24* and *KLF2,* had a wider gene expression for all the tissues explored in the *GTEx* v8 dataset. *CSF3* had an increased expression level in adipose tissue, fibroblasts, the pituitary, the prostate, the fallopian tubes, and musculoskeletal tissue. *GSMDA* expressions were significant only in two tissues, both relating to skin. Other genes, *CHERP, KLF2, MED24, PSMD3,* and *THRA,* showed heightened expression levels in an overlapping manner for a wide variety of tissues. Significant overlap in gene enrichment was seen in chromosomes 17 and 19 (Supplementary Figure 16). In the gene set analysis, for the 10 leading pathways, only the potassium ion transport pathway remained significant after applying the Bonferroni correction (Supplementary Table 5).

We also explored the network of genes closest to the significant SNPs in the trans-ancestry meta-analysis (Supplementary Figure 17). The network analysis of significant genes in the trans-ancestry meta-analysis revealed evidence of high co-expression (56.34%), physical interaction (25.35%), and common pathways (13.44%). For the construction of the best-fit PRS, we used 74,377 of 55,333,12 variants using the clumping and thresholding (C+T) method for SNP thinning (Supplementary Figures 18-19). The strata plot indicates a dose-response relationship between allele scores and the phenotype, although this was not discernible in the two uppermost quantiles (Figure 2). The gender-wise distribution of NLR and PRS is shown in Supplementary Figure 20.

**Figure 2.**
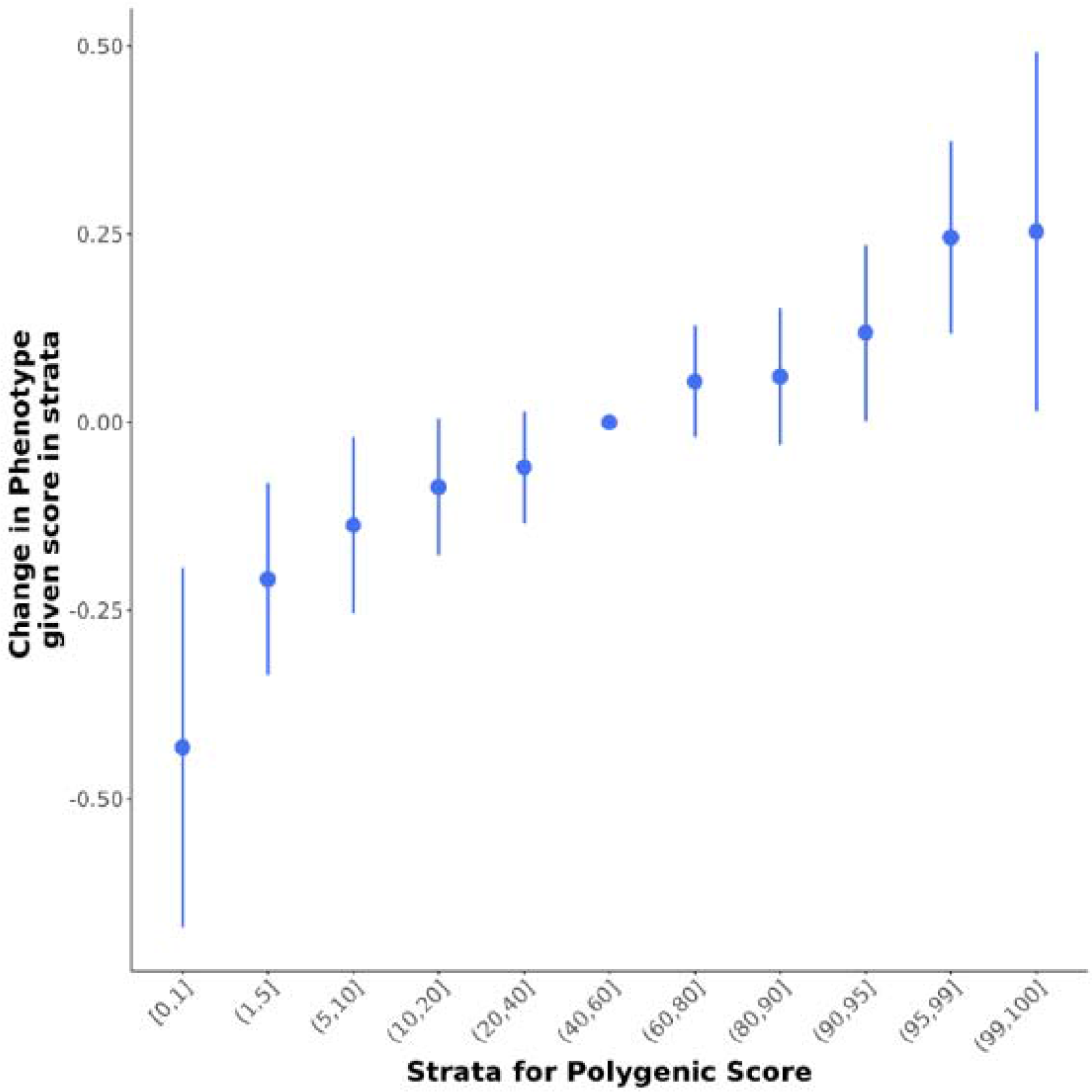
Strata Plot for the NLR PRS in SHARE. Note: The plot illustrates the change in the computed PRS for NLR on the x-axis and the values of NLR on the y-axis. The blue dot represents the point estimates, and the lines represent the upper and lower 95% confidence intervals for the change in NLR corresponding to the PRS. In general, the NLR phenotype increases with the predicted PRS scores, indicating that PRS can adequately explain the individual variation of the NLR.

### Competing risks analyses

In the SHARE cohort (n=3,892), the Fine and Gray model did not indicate any statistically significant association with DR (n=1,610) in both adjusted and unadjusted models (Supplementary Table 6). There were 184 competing events, and the mean follow-up time was 4.8 years. The adjusted subhazards for NLR PRS with DR were 1.01 (95% CI: 0.97, 1.06, p=0.48). Thus, PRS for NLR may not be useful for predicting the long-term manifestation of diabetic retinopathy.

### Mendelian randomization analysis for DR using NLR allele scores

The regression coefficient between the phenotype and PRS was statistically significant at 0.11 (p=3.41×10^-15^) (Supplementary Figure 21). The PRS predicted only 1.2% of the NLR variation, which improved to 3.5% after the addition of age and sex (Supplementary Table 7). We analysed 3,561 Scottish individuals (Supplementary Figures 22-23) with data on PRS, NLR, and diabetic retinopathy for the MR analysis, of which 1,537 (43.1%) had DR. The F-statistic indicates a strong relationship between the PRS and NLR (close to 60), even after adjusting for the effects of age and sex. The results from the MR analysis are given in Figure 3. The estimates were very similar between the models. Logistic-linear and TSPS estimated a non-significant effect (n= 3,561, beta = 0.22, 95% CI: - 0.25, 0.70, p=0.35). For the Cox IV method (n=3,081, DR=1216), The mean follow-up time was 4.8 years. Again, there were no statistically relevant findings between a unit change in the PRS and hazards for DR (-0.15, 95% CI: -0.65, 0.34, p=0.55). Overall, our analysis provided no evidence of a causal association between genetically determined levels of NLR and DR.

**Figure 3.**
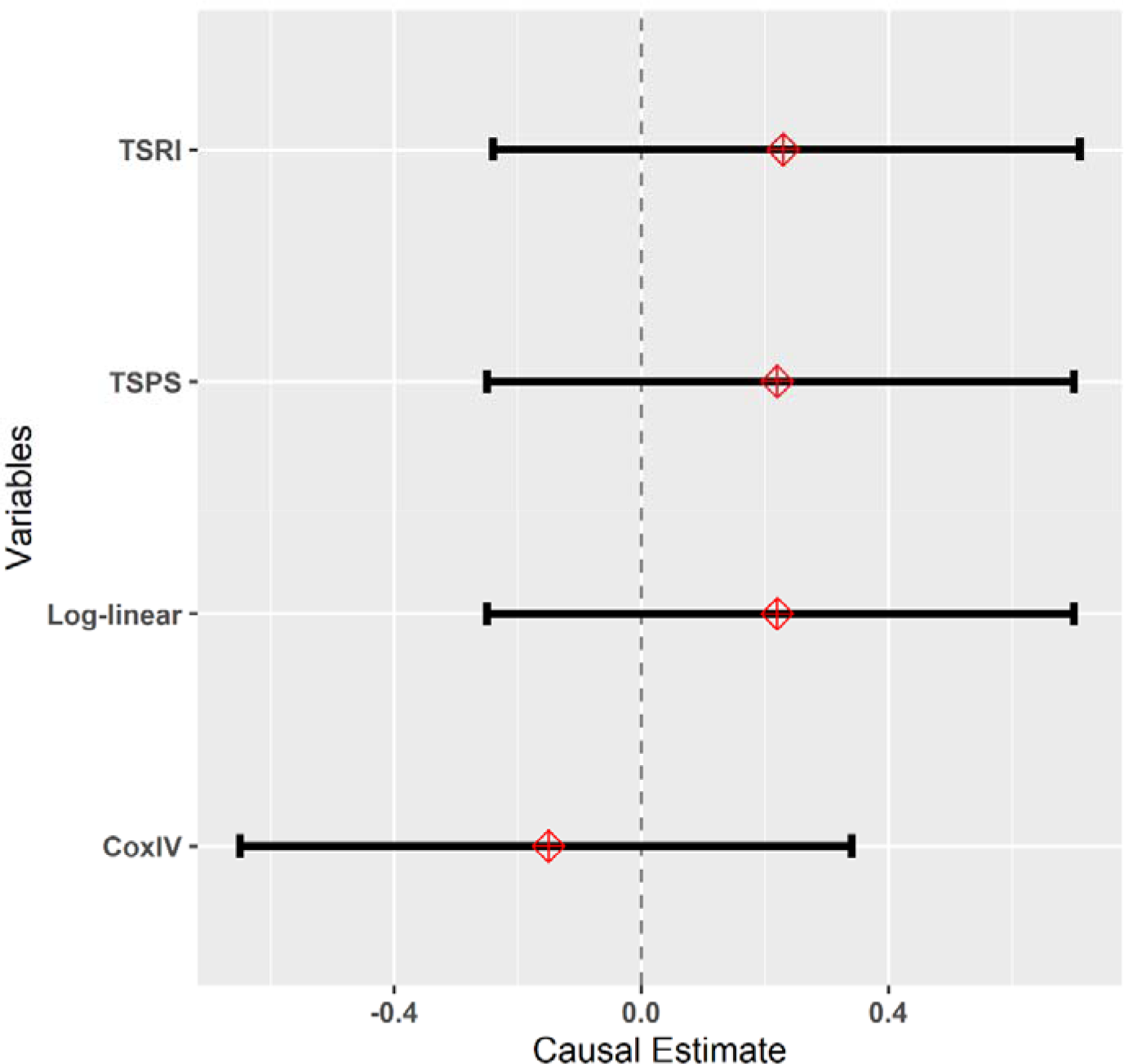
MR analysis of NLR allele score with any DR. Note. Log-linear Logistic linear, TSRI two-stage residual inclusion, TSPS two-stage predictor substitution, IV Instrumental Variable, SE standard error, CI confidence interval

## Discussion

Our study found a divergence in the genetic architecture of NLR between South Asians and white Europeans. South Asian populations are generally underrepresented in genetic studies of NLR, and the present results may cover this gap. In general, white Europeans had a much higher NLR relative to other ancestries.^41^ The heritability estimates (h^2^) in our work also reflect this disparity. Our heritability estimates almost matched those in the previous Dutch twins’ study.^42^ White blood cells are genetically determined; an example is the Duffy-Null polymorphism among the African population arising due to selection pressures.^43^

Remarkably, NLR was significantly correlated with multiple inflammatory markers such as CRP and IL6, but not platelet-lymphocyte ratio (PLR), implying that the variation of WBC ratios could provide biological insights.^42^ Overall, some important genes are associated with white blood cell-related traits, but not specifically with the NLR. Chromosome 17q21 contains many genes that exert substantial pleiotropic effects across different WBC subcomponents and is highly expressed in inflammatory disorders such as asthma and diabetes. ^44,45^ Given diabetes and inflammation, there was evidence of *PSMD3* involvement through insulin resistance and receptor tyrosine kinase (RET) signalling, respectively.^46–49^ *CSF3*, *MED24*, and *PSMD3* genes were reported together to influence neutrophil counts in a Bayesian meta-analysis involving multiple ancestries.^50^ Similarly, *GSDMA*, *THRA*, *EPS15L1*, *CALR3*, *CHERP*, and *MED24* were also associated with multiple white blood cell types, including neutrophils and lymphocytes.^45^ Importantly, the rest of the significant genes; *CTC429P9.4*, *CTD3222D19.2,* and *SLC35E1,* do not seem to have a direct link with white blood cell physiology and are therefore novel findings that merit further mechanistic studies. Another novel NLR gene was *CLECL1*, which takes part in the T lymphocyte pathway but is also differentially expressed in clusters of inflammatory conditions such as diabetes mellitus and dyslipidemia.^51^

We replicated rs6841652 of the *LINCO2513/TBC1D gene*. This SNP was also associated with monocyte-lymphocyte ratio and multiple SNPs, that could be traced back to the same genes were significant for NLR.^52^ We were also able to detect associations with the thyroid hormone receptor gene (*THRA*) that were previously reported for NLR in the Korean population.^53^ The complex cross-talk between thyroid hormones and immune cells is vital for the proper regulation of inflammatory activity.^54^ Most genes from the gene-based set were related in varying degrees, as elicited in the network analysis. Moreover, the gene set analysis overwhelmingly showed affinity for the potassium ion transportation pathway.

One of the strengths of the study is that, we did not transform the NLR as opposed to previous studies ^53,55^ As data transformation could rasie concerns of reduced statistical power and ease of beta interpretation.^56^ A summarized NLR could override possible bias arising from health conditions, outliers, and other unusual deviations from the original phenotype for accurate genome-wide scanning. We further refined the SNP results in each ancestry to identify independent SNPs free from LD. To our knowledge, this is the first study that details the generation of an allelic summary score for NLR and uses it for a causal analysis for DR. We could not replicate the NLR association for DR in the observational study using the PRS.^57–59^ The high F statistics for the IV-phenotype relation do not completely eliminate the possibility of bias.^60^ It is also possible that the NLR is regulated at a more downstream level by several competing inflammatory pathways or environmental stimuli. Therefore, the current findings may not be sufficient to rule out the role of inflammation in DR. The major limitation was that a substantial proportion of the dataset was used to train the PRSice-2 algorithm, leaving a much smaller sample to investigate DR. Due to ancestry differences, PRS and subsequent analysis were conducted only in the Scottish cohorts. Finally, our one sample MR analysis is relatively less powerful compared to a two-sample MR.^61,62^ Additional studies are required to understand the genetic mechanisms influencing the NLR between ancestrally diverse populations and subpopulations.

## Conclusion

In summary, we report multiple novel intergenic SNPs associated with the NLR in addition to those identified for WBC previously and most of the identified SNPs were in the intergenic regions. There was no evidence to suggest causality between systemic inflammation represented by NLR and DR. In the context of chronic diseases, our findings may provide insights into inflammatory pathways to develop better strategies to deal with inflammatory outcomes.

## Supporting information

STREGA Check List

## Data Availability

All data produced in the present study are available upon reasonable request to the authors.

## Availability of data and materials

The Scottish and DMDSC datasets are stored electronically in a secure environment. The data is not currently available in the public domain as the participant consent restricts the public sharing of the data. Researchers will require prior approval from the HIC and DMDSC for accessing the related datasets.

## Abbreviations

BMI: Body Mass Index
CI: Confidence Interval
CIF: Cumulative Incidence Function
chr: Position: Chromosome to Position
CURES: Chennai Urban Rural Epidemiology Study
DBP: Diastolic Blood Pressure
DMDSC: Dr. Mohan’s Diabetes Specialties Clinics
DR: Diabetic Retinopathy
eGFR: Estimated Glomerular Filtration Rate
FE: Fixed Effects
FGR: Fine and Gray model
FUMA: Functional Mapping aand Annotation of Genome-wide Association Studies
GCTA: Genome-wide Complex Trait Analysis
GIF: Genomic Inflation Factor
GoDARTS: Genetics of Diabetes Audit and Research in Tayside Scotland
HbA1c: Glycated Haemoglobin
HDLc: High-density lipoprotein cholesterol
HIC: Health Informatics Centre
HRC: Haplotype Reference Consortium
HWE: Hardy-Weinberg Equilibrium
LD: Linkage Disequillibrium
LDL-C: Low-density Lipoprotein -Cholesterol
MAGMA: Multi-marker Analysis of GenoMic Annotation
MDRF: Madras Diabetes Research Foundation
NLR: Neutrophil Lymphocyte Ratio
Non-HDL-C: Non–high-density Lipoprotein Cholesterol
PLR: Platelet-lymphocyte ratio
PRS: Polygenic Risk Score
RET: Receptor Tyrosine Kinase
SBP: Systolic Blood Pressure
SD: Standard Deviation
SHARE: Scottish Health Research Register
SE: Standard Error
sHR: Sub Hazard Ratio
SNP: Single Nucleotide Polymorphisms

## Acknowledgments

The authors would like to thank all participants in the GoDARTS, SHARE, and DMSDC cohorts. We would also like to thank the Health Informatics Centre (HIC), University of Dundee for providing access to the Scottish Data and staff of MDRF, Chennai.

## Author Contributions

C.N.A.P. and A.L.R were involved in the conceptualisation, analysis, interpretation and manuscript writing. A.T.N., M.C., and C.N. were involved in GWAS analysis and heritability estimation. SS made a substantial contribution toward ensuring data quality and manuscript revision. M.K.S, V.R., R.M.A., W.M. and V.M. revised the manuscript critically for important intellectual content and provided guidance for data analysis. All authors provided approval for manuscript publication. C.N.A.P. is the guarantor of this work, had access to the database, and takes full responsibility for the integrity of the data, decision to submit and publish the manuscript.

## Supplemental Material

Supplementary File 1 – Figures

Supplementary File 2 – Tables

STREGA check list

