## Supplementary Figures for "Multi-ancestry genome-wide association study of neutrophil-lymphocyte ratio and polygenic risk score development to explore causal association with diabetic retinopathy"

**SUPPLEMENTARY DATA**

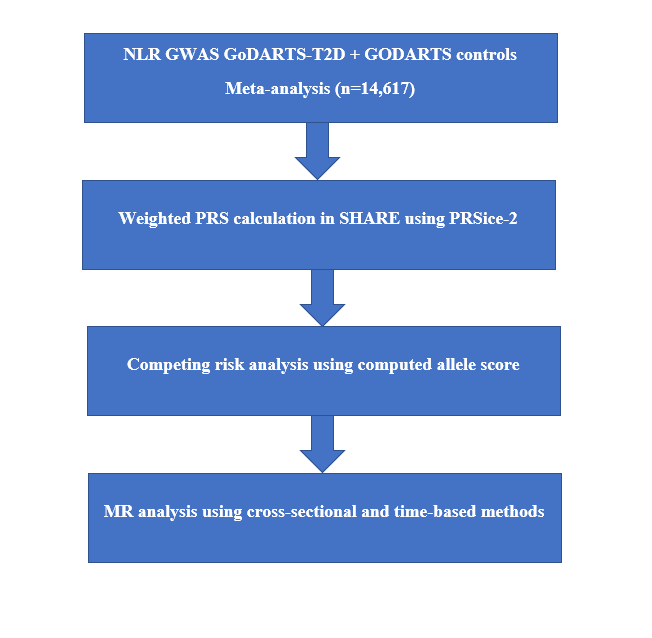

Supplementary Figure 1. Workflow Schema depicting the PRS computation for NLR and subsequent analysis

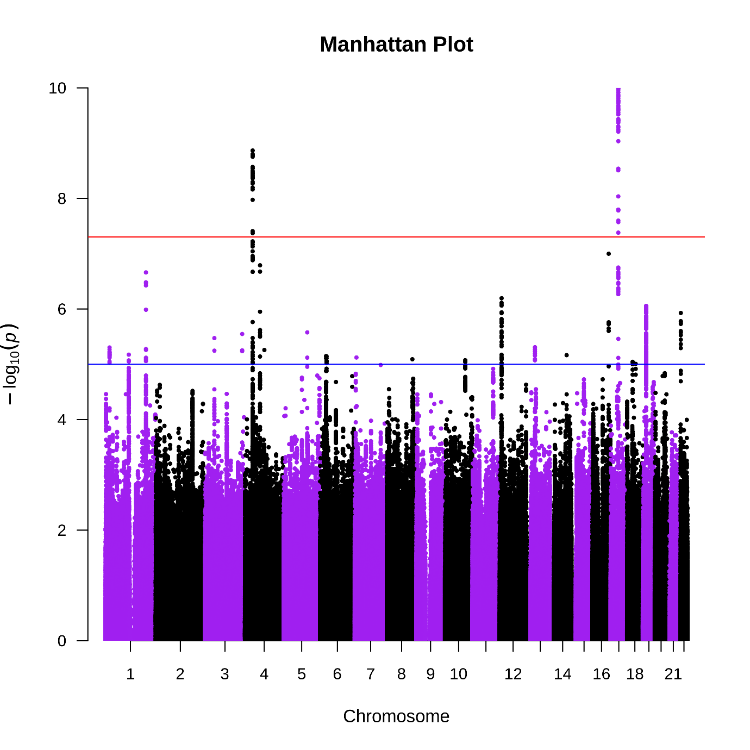

Supplementary Figure 2. Manhattan plot showing statistically significant SNPs from the fixed effects meta-analysis associated with NLR in the Scottish cohort (n= 21,153)

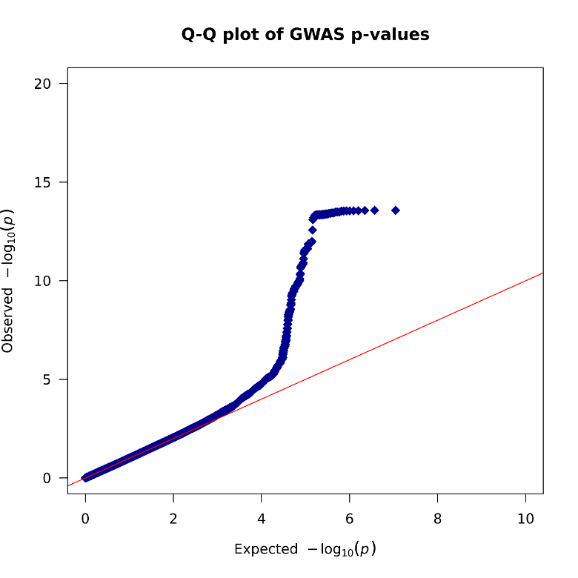

Supplementary Figure 3. Q-Q plot from the fixed effects meta-analysis from the GWAS NLR in the Scottish cohort (n= 21,153, λ= 1.09)

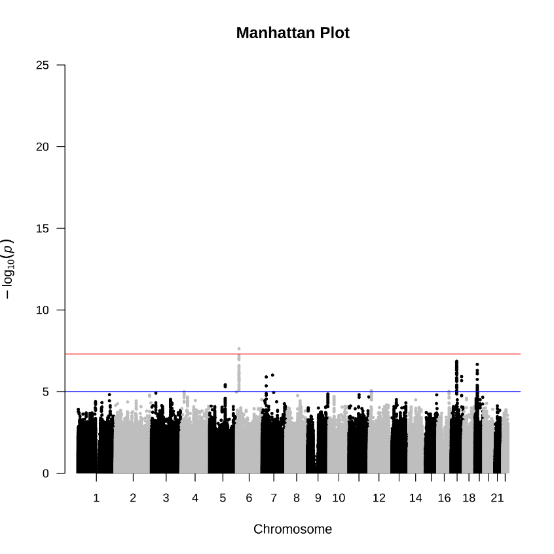

Supplementary Figure 4. Manhattan plot showing statistically significant SNPs from the fixed effects meta-analysis associated with NLR in the DMDC cohort (n= 8,164)

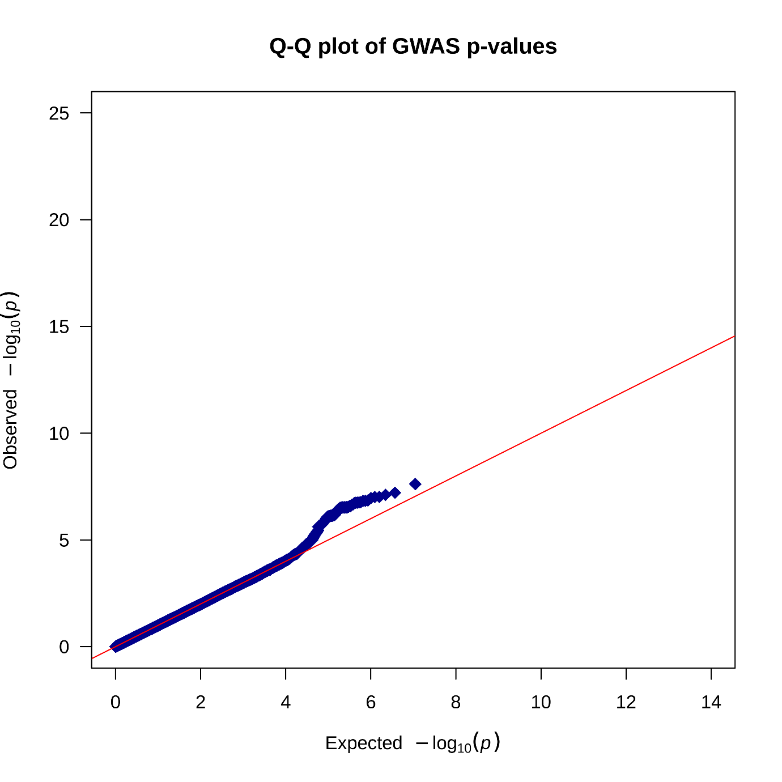

Supplementary Figure 5. Q-Q plot from the fixed effects meta-analysis from the GWAS NLR in the DMDC cohort (n= 8,164, λ=1.01)

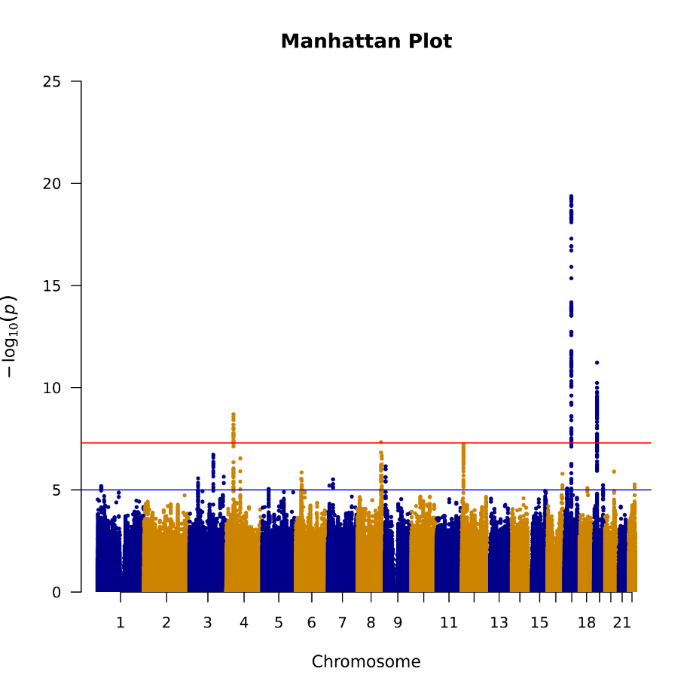

Supplementary Figure 6. Manhattan plot showing statistically significant regions across chromosomes from the fixed effects meta-analysis associated with NLR in the Scottish and Indian cohorts (n= 29,317)

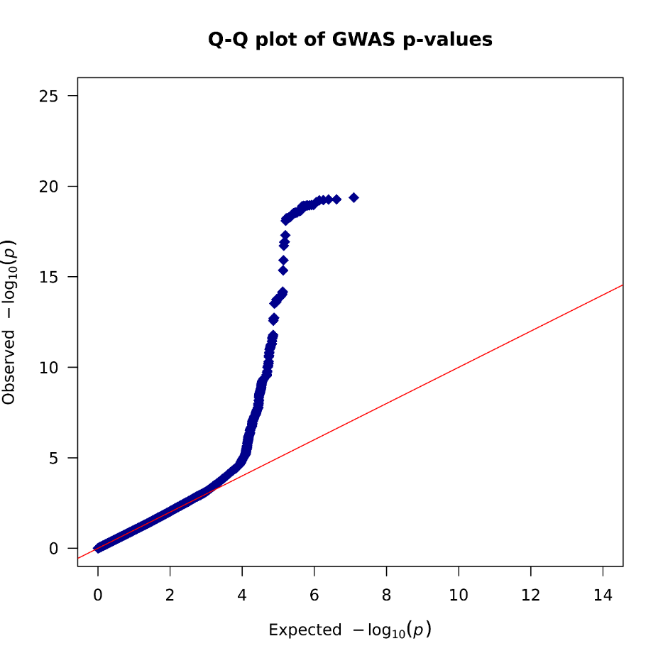

Supplementary Figure 7. QQ plot showing statistically significant regions across chromosomes from the fixed effects meta-analysis associated with NLR in the Scottish and Indian cohorts (n= 29,317, λ=1.07)

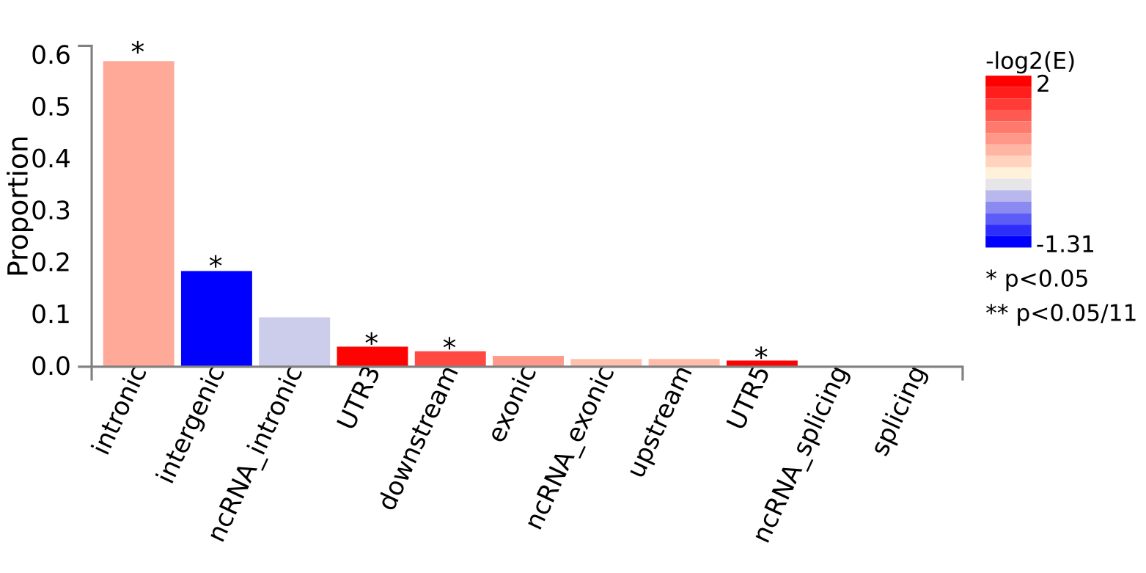

Supplementary Figure 8. Functional consequences of SNPs on genes

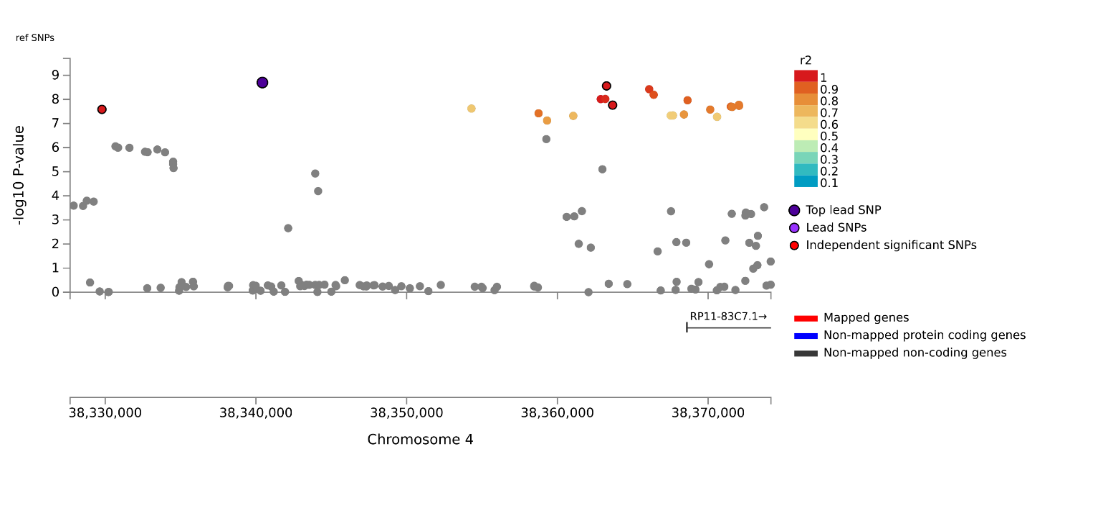

Supplementary Figure 9. Regional plot visualizing the significant SNPs on Chromosome 4

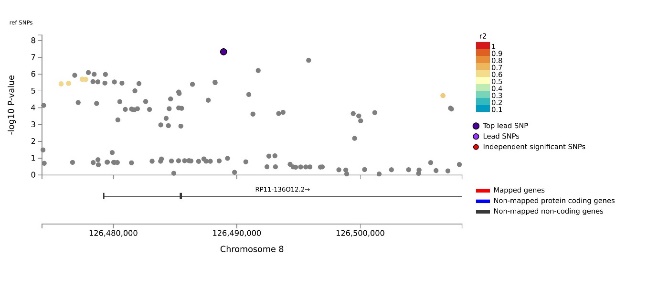

Supplementary Figure 10. Regional plot visualizing the significant SNPs on Chromosome 8

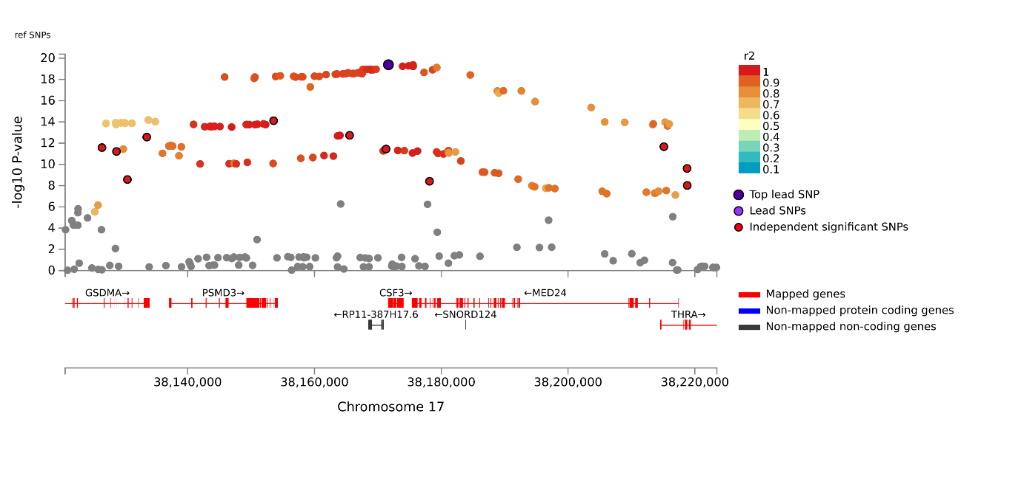

Supplementary Figure 11. Regional plot visualizing the significant SNPs on Chromosome 17

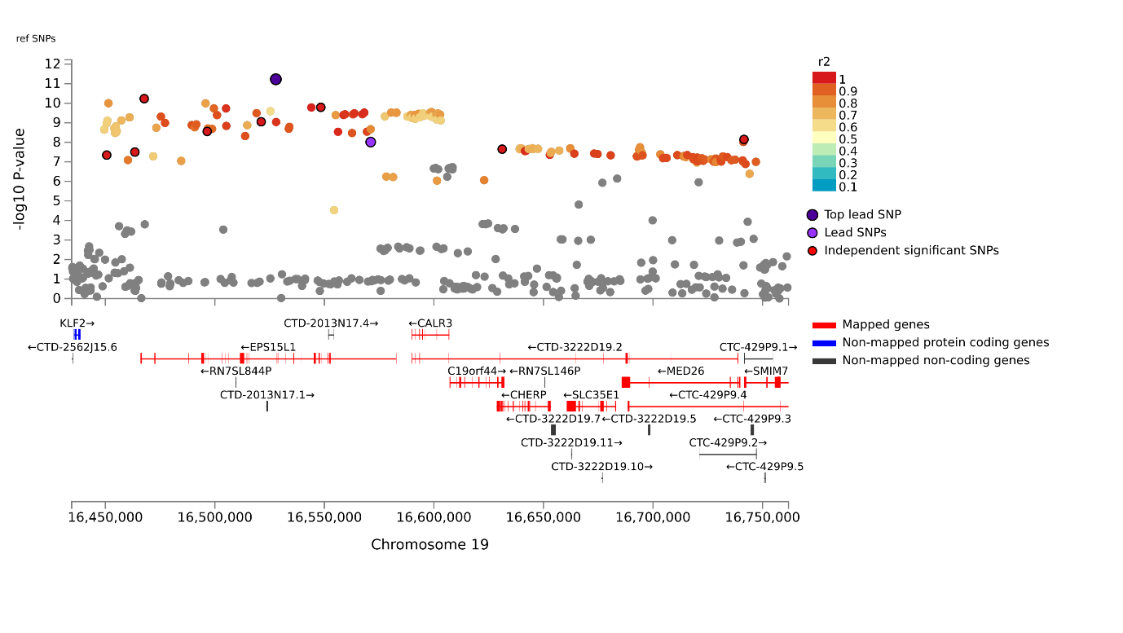
 Supplementary Figure 12. Regional plot visualizing the significant SNPs on Chromosome 19

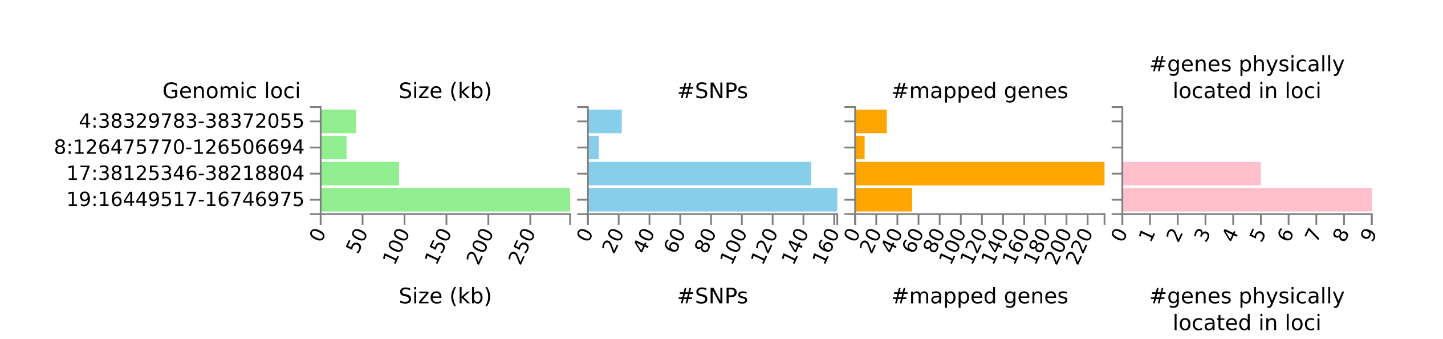

Supplementary Figure 13. Summary of risk loci and genes associated with GWAS of NLR in the trans-ancestry meta-analysis

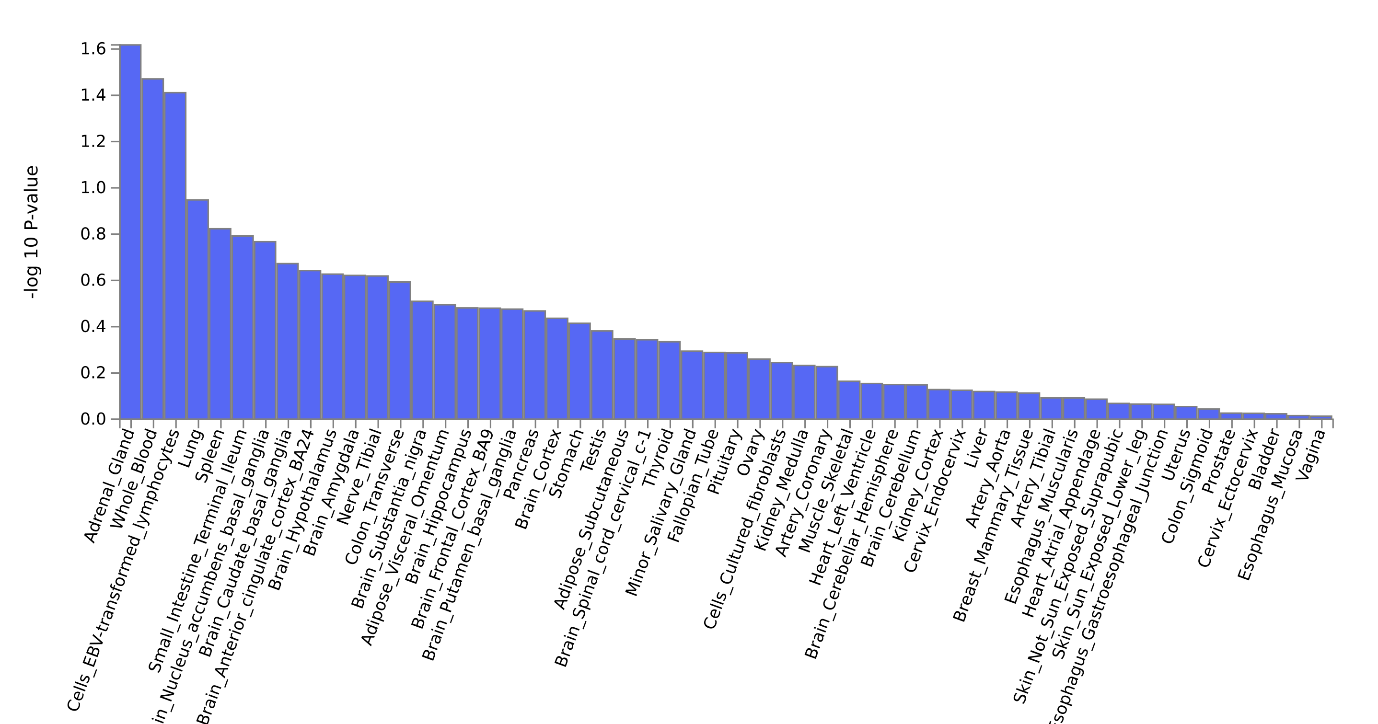

Supplementary Figure 14. Differential expression of protein coding genes from the trans-ancestry meta-analysis of Scottish and Indian cohorts measured in the GTEx dataset. Note. Blue rising blocks indicate that the genes were generally upregulated in the tissues.

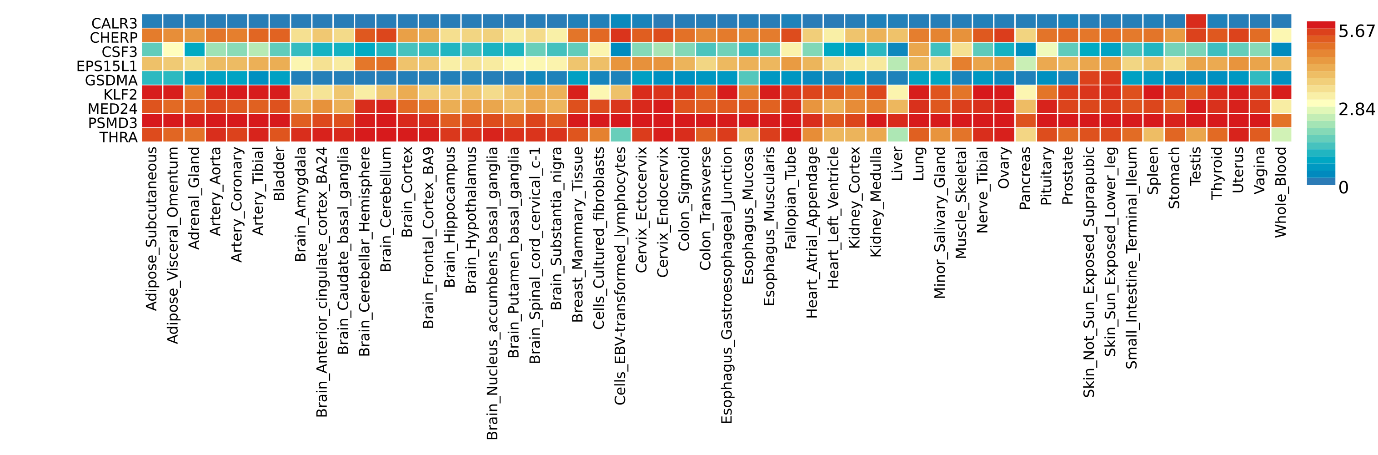

Supplementary Figure 15. Gene expression heatmap by specific tissue type in the GTEx data for the significant protein coding genes from the gene-based analysis

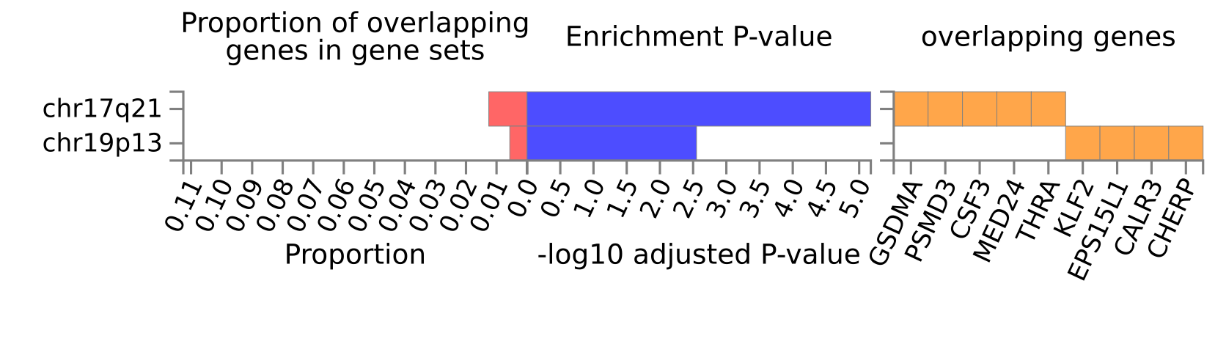
 Supplementary Figure 16. Proportion of overlapping gene set in chromosome 17 and chromosome 19

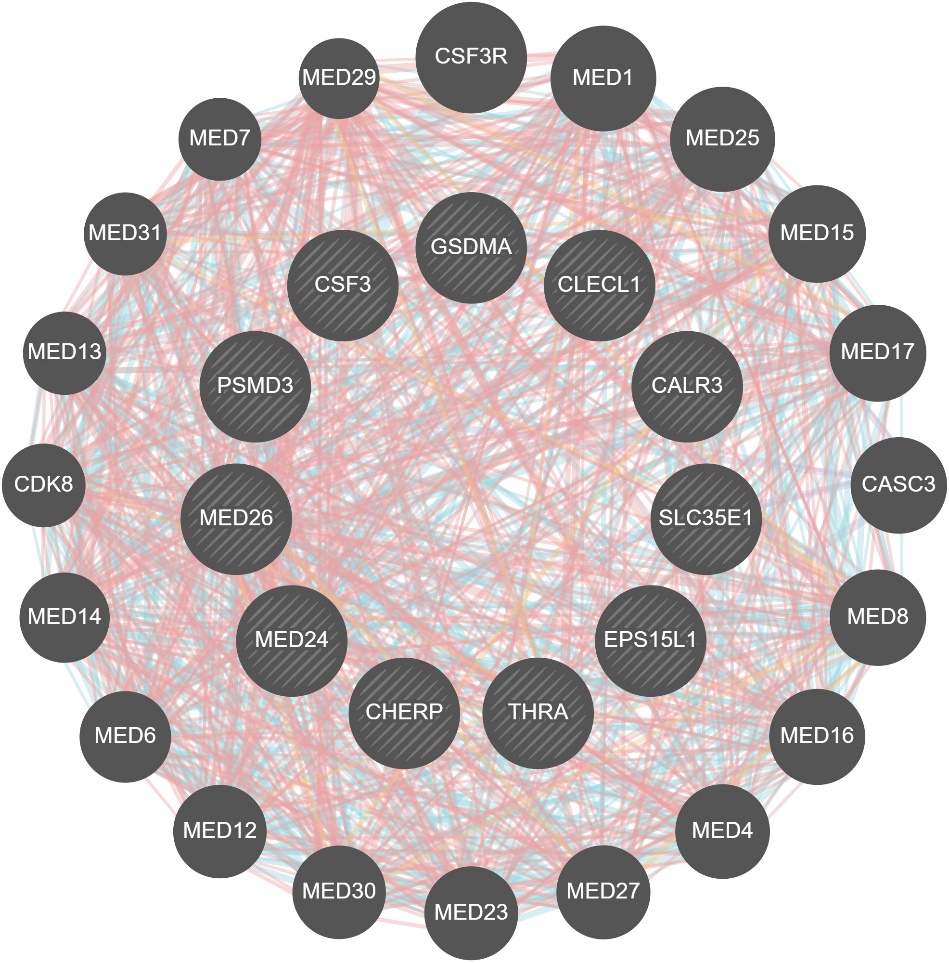

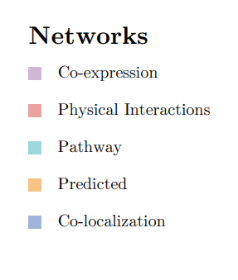

Supplementary Figure 17. Visualization of gene networks illustrating the relationships and enrichment pathways for major genes influencing NLR phenotype identified from the gene-set analysis

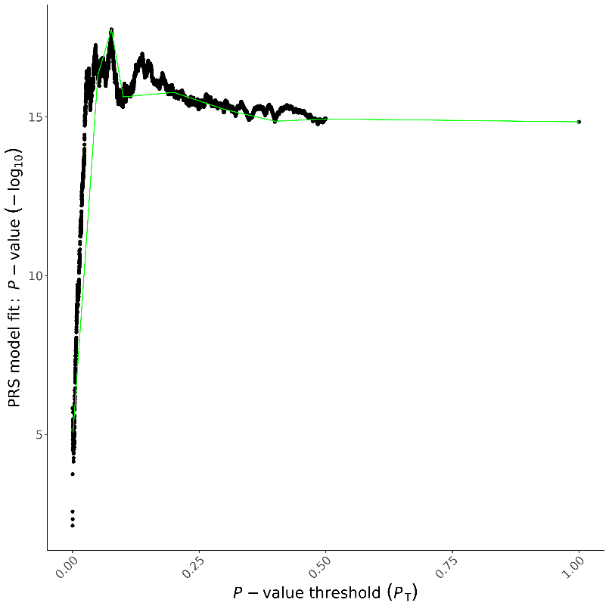

Supplementary Figure 18. Model results of the best fit PRS analysis

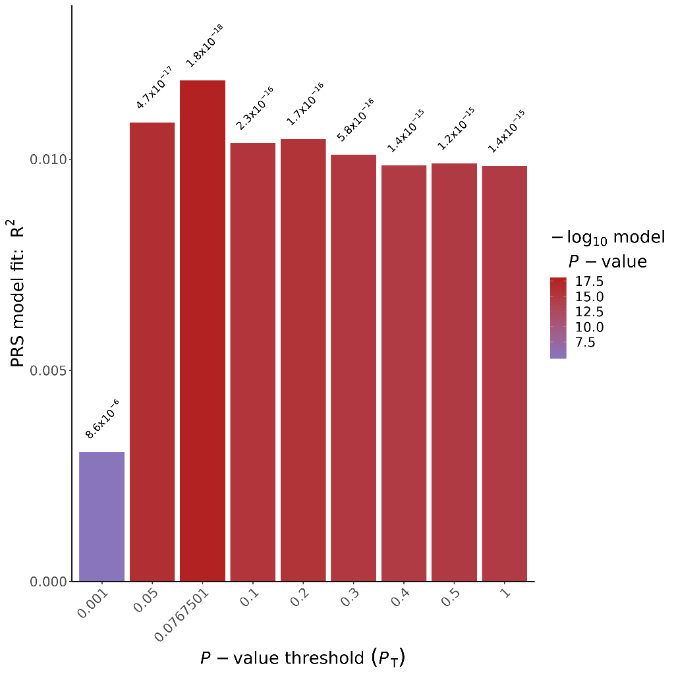

Supplementary Figure 19. Plot showing the model fit R square for different p-value thresholds

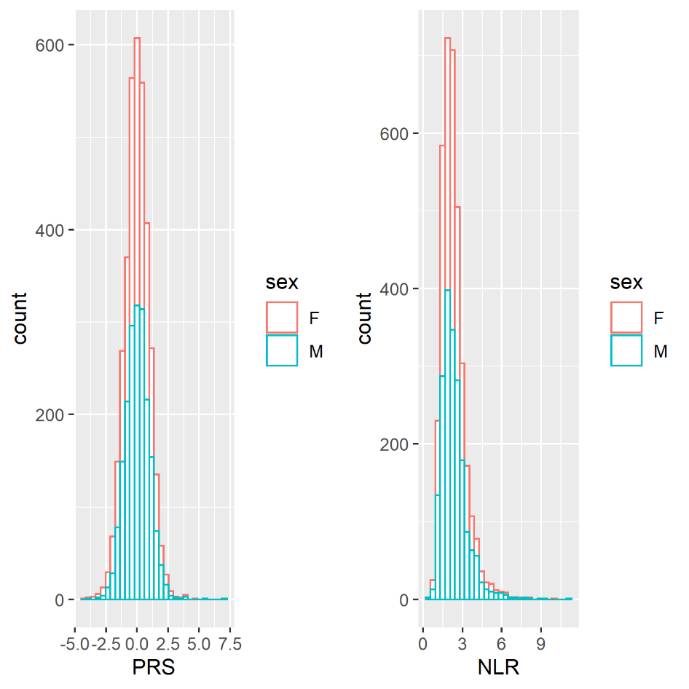

Supplementary Figure 20. NLR and estimated NLR PRS distribution stratified by gender in the SHARE.

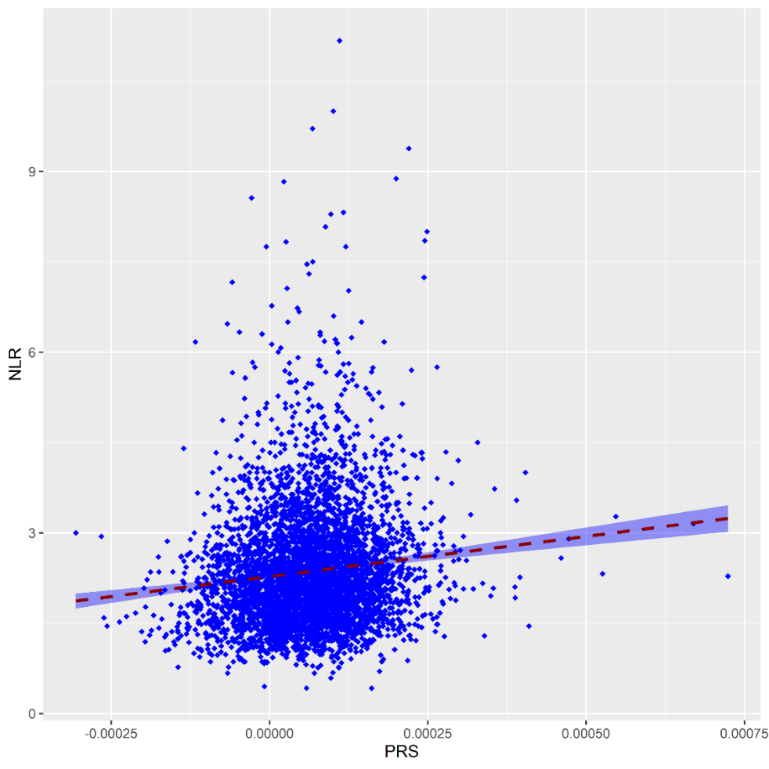

Supplementary Figure 21. Correlation between the NLR phenotype with PRS for SHARE cohort

DR Screening with complete covariate information (n=10,414)

6,522 participants were excluded:

Linked PRS data (n=7,034)

No recorded DR grade (45,5records, 66,197 PROCHI)

First screening visit >=1990-01-01 (43,6440 PROCHI, 64,879 PROCHI)

No DM diagnosis date -55728

Date out < DM diagnosis date:55616

Death date > DM diagnosis date:55612

Follow-up date > T2D date (55,327 PROCHI)

Combined data with NLR PRS, DR screening and covariates (n=3,892)

331 participants were excluded:

Linked NLR phenotypic data (n=6,681) with PRS, DR and covariates (n=3,662), Missing NLR values excluded (n=101)

Missing covariat

Final MR analysis sample (n=3,561)

Supplementary Figure 22. Flow diagram showing sample selection for MR in the cross-sectional analysis

DR Screening with complete covariate information (n=10,414)

6,522 participants were excluded:

Linked PRS data (n=7,034)

No recorded DR grade (45,5records, 66,197 PROCHI)

First screening visit >=1990-01-01 (43,6440 PROCHI, 64,879 PROCHI)

No DM diagnosis date -55728

Date out < DM diagnosis date:55616

Death date > DM diagnosis date:55612

Follow-up date > T2D date (55,327 PROCHI)

Combined data with NLR PRS, DR screening and covariates (n=3,892)

811 participants were excluded:

Linked NLR phenotypic data (n=11,397) with PRS, DR and covariates Missing covariate

Final Cox IV MR analysis sample (n=3,081)

Supplementary Figure 23. Flow diagram showing sample selection for the Cox MR analysis
