## Supplementary Tables for "Multi-ancestry genome-wide association study of neutrophil-lymphocyte ratio and polygenic risk score development to explore causal association with diabetic retinopathy"

Supplementary Table 1. Cohort wise description of participant characteristics for the various Scottish cohorts used for the discovery GWAS (n= 21,153)

| Cohort | Sample  Size | Median NLR (IQR) | Median Age (IQR) | Sex (M %) | DM (%) | GIF ( λ) |
| --- | --- | --- | --- | --- | --- | --- |
| **GoDARTS** | | | | | | |
| Affymetrix | 3,754 | 2.30  (1.83-2.93) | 74.75  (67.01-81.43) | 2,001 (53.3%) | 3,651 (97.2%) | 1.02 |
| Illumina | 3,549 | 2.29  (1.75-2.94) | 74.19  (65.43- 81.42) | 2,035(57.3%) | 3,417(96.3%) | 1.04 |
| Broad | 1,834 | 2.15  (1.68-2.70) | 70.12  (59.03-79.42) | 924 (50.4%) | 951(51.8%) | 0.99 |
| SHARE | 6,446 | 2.14  (1.70-2.73) | 66.49  (56.28-75.30) | 3,401 (52.8%) | 4,562 (70.7%) | 1.01 |
| GODARTS controls | 5,570 | 2.04  (1.62-2.71) | 69.39  (58.72 -79.75) | 2,727 (48.9%) | 433 (7.7%) | 1.02 |
| GoDARTS+ SHARE  + GoDARTS controls | 21,153 | - | - | -- | -- | 1.09 |
| DM diabetes mellitus, GoDARTS cohort consists of 3 genotypic platforms – Affymetrix, Illumina and Broad, GoDARTS controls, SHARE Scottish Health Research Register, IQR Interquartile Range, GIF Genomic Inflation Factor | | | | | | |

Supplementary Table 2. Overall description of the DMDSC cohorts (n=8,164)

| Cohort | Sample  Size | Median NLR (IQR) | Median Age (IQR) | Sex (M %) | DM (%) | GIF ( λ) |
| --- | --- | --- | --- | --- | --- | --- |
| DMDSC cohort 1 | 5,773 | 1.93 (1.54-2.39) | 58.16 (47.28-66.44) | 3489(60.0) | 5,737 (99.37) | 1.03 |
| DMDSC cohort 2 | 2,391 | 1.90 (1.53-2.40) | 53.82 (41.23-63.98) | 1445(60.4) | 2,140 (89.5) | 1.01 |
| Combined | 8,164 |  |  |  |  | 1.01 |
| DM diabetes mellitus, DMDSC Dr. Mohan’s Diabetes Specialties Centre, IQR Interquartile Range, GIF Genomic Inflation Factor | | | | | | |

Supplementary Table 3. Statistically significant SNPs identified in the FE metanalysis of DMDSC cohorts (n=8,164)

| **rsid** | **chr: Pos** | **Genotype** | **MAF** | **Beta** | **p-value** | **Location** | **Gene** |
| --- | --- | --- | --- | --- | --- | --- | --- |
| rs17330192 | 6: 17589375 | T/C | 0.34 | -0.11 | 1.8x10^-8^ | Intron | LOC102724591 |
| rs139801819 | 6:17588945 | G/A | 0.34 | -0.11 | 2.4 x10^-8^ | Intron | LOC102724591 |

Note. MAF minor allele frequency, chr: pos chromosome to position

Supplementary Table 4. Estimates of SNPs from the trans-ancestry GWAS independently associated with NLR in the discovery cohort (n=29,317)

| **Rsid** | **chr: Position** | **Genotype** | **MAF** | **Beta** | **P** | **I^2^** | **Location** | **Nearest Genes** |
| --- | --- | --- | --- | --- | --- | --- | --- | --- |
| rs3826331 | 17:38150492 | T/C | 0.46 | -0.07 | 8.05x10^-19^ | 0.19 | Intron | PSMD3 |
| rs6841652 | 4: 38363262 | T/C | 0.39 | -0.05 | 2.76x10^-9^ | 0.47 | - | LINCO2513, TBC1D1 |
| rs16850400 | 4: 74927915 | T/C | 0.45 | 0.04 | 1.24x10^-6^ | 0.42 | - | - |

MAF minor allele frequency, chr: Position chromosome to position, I^2^ heterogeneity index indicating the percentage of variation across studies

Supplementary Table 5. Results from the gene-set analysis generated using MAGMA for the GWAS summary in the fixed effects trans-ancestry meta-analysis

| **Gene Set** | **N genes** | **Beta** | **SD** | **SE** | **P** | **P_bon_** |
| --- | --- | --- | --- | --- | --- | --- |
| Potassium ion transport | 111 | 0.40 | 0.043 | 0.09 | 3.0x10^-6^ | 0.04 |
| Cellular potassium ion homeostasis | 7 | 1.52 | 0.041 | 0.37 | 1.8x10^-5^ | 0.27 |
| Potassium ion transmembrane transporter activity | 79 | 0.41 | 0.037 | 0.10 | 7.2x10^-5^ | 1 |
| Reactome FCGR activation | 4 | 1.54 | 0.031 | 0.43 | 0.0001 | 1 |
| NUMATA_CSF3_SIGNALING_VIA_STAT3 | 14 | 0.89 | 0.034 | 0.25 | 0.0002 | 1 |
| Nakayama_soft_tissue_tumors_PCA1_DN | 41 | 0.49 | 0.032 | 0.14 | 0.0002 | 1 |
| Calcium activated potassium channel activity | 9 | 1.16 | 0.035 | 0.34 | 0.0003 | 1 |
| Cellular monovalent inorganic cation homeostasis | 44 | 0.45 | 0.030 | 0.13 | 0.0005 | 1 |
| Curated gene sets: liu targets of_vMyb vs_cMyb_up | 6 | 0.82 | 0.020 | 0.25 | 0.0005 | 1 |
| Plasma membrane protein complex | 270 | 0.18 | 0.030 | 0.05 | 0.0005 | 1 |

Supplementary Table 6. Fine and Gray analysis of NLR PRS with DR incidence (n=3,892)

| **Variable** | **sHR (95%CI)** | **adj. sHR (95% CI)** | **p** |
| --- | --- | --- | --- |
| NLR PRS | 1.02(0.96, 1.07) | 1.01 (0.97,1.06) | 0.48 |
| HbA1c | - | 1.15 (1.12, 1.18) | <0.001*** |
| Age | - | 0.99 (0.99,1.00) | 0.07 |
| Sex (M) | - | 1.05 (0.95,1.16) | 0.29 |
| BMI | - | 0.98 (0.98,0.99) | <0.01** |
| Creatinine | - | 1.004 (1.002,1.005) | <0.001*** |

Note. NLR, Neutrophil–Lymphocyte Ratio. PRS, Polygenic risk score. sHR, Subhazard ratio. ^**^p<0.01; ^***^p<0.001. 95% CI, 95% Confidence Interval.

Supplementary Table 7. Association of the NLR PRS with the phenotype and corresponding adjusted r square and F-statistics

| **Variable** | **Estimate** | **SE** | **p** | **Adjusted R-squared** | **F-statistic** |
| --- | --- | --- | --- | --- | --- |
| PRS (crude) | 1330.67 | 168.42 | <0.001*** | 0.012 | 62.42 |
| PRS (Adjusted) | 1255.79 | 166.68 | <0.001*** | 0.035 | 59.99 |

Note. PRS adjusted for age and sex. ^***^p<0.001. SE Standard Error
